# Cervical Cancer Screening Seeking Behavior among Female Community Health Volunteers of Surkhet District

**DOI:** 10.1101/2024.07.25.24311012

**Authors:** Yamuna Thapa, Bimala Bhatta

## Abstract

**Background:** Human Papilloma virus (HPV) associated cervical cancer is the fourth most common cancer in women worldwide and it is the leading cause of death among women in Nepal. The screening seeking behavior encompasses a woman’s decision to engage in medical evaluations essential for the early detection and prophylaxis of cervical cancer. Even though there is a proven importance of cervical cancer screening, the incidence and mortality rate in Nepal is high. Therefore, we aimed to assess the cervical cancer screening seeking behavior and its associated factors among female community health volunteers (FCHVs) of Surkhet district, Nepal.

**Methods:** A community based cross-sectional study was carried out in the municipalities and rural municipalities of Surkhet district. A pre-tested structured interview was conducted among 148 FCHVs from 30-49 years.

**Results:** This study showed that 90 (60.8%) of FCHVs have cervical cancer screening seeking behavior. Age (AOR: 7.2, 95% CI: 3.01-17.3) and marital status (AOR: 9.2, 95% CI: 2.6-166.2) of FCHVs were significant factors for cervical cancer seeking behavior.

**Conclusion:** These findings highlight the importance of demographic factors in promoting screening participation among FCHVs. Enhancing cervical cancer screening rates among FCHVs requires multifaceted approaches that address both individual perceptions and barriers. Interventions should focus on increasing accessibility, improving education and awareness programs, and providing tailored support to different demographic groups.

## Introduction

Cervical cancer screening seeking behavior refers to a woman’s decision to undergo medical test essential for early detection and prevention of risk. The cervical cancer screening aims to detect precancerous lesions in cervix, which can prevent the invasion of these cells and treat them earlier. This ultimately reduces the incidence and mortality rate of cervical cancer[1,2]. Women should have screenings every 5 years after the age of 30 and those living with HIV every 3 years from age 25 years. Cervical cancer screening methods include VIA, VILI, colposcopy, cervical biopsy, Pap smear, and HPV testing. The screening is influenced by awareness, beliefs, access, financial constraints, and healthcare infrastructure. However, LMICs often face resource and skilled professional shortages for screening. Therefore WHO and ACCP advocate VIA as a cost-effective method for screening. [3].

Despite being both a preventable and potentially curable disease, cervical cancer is the fourth most common in women worldwide, with 6,04,000 new cases and 3,42,000 deaths annually. The highest rates of cervical cancer incidence and mortality are in sub-Saharan Africa (SSA), Central America and South-East Asia [4]. Almost all cervical cancer cases (99%) are linked to infection with high-risk human papillomaviruses (HPV), an extremely common virus transmitted through sexual contact[5]. HPV is not solely sufficient to cause cervical cancer. In addition, with HPV, other variables like parity, oral contraceptive use, smoking, early marriage, multiple sexual partners, poor genital hygiene, week immune system, STDs/HIV and poor nutrition contribute to the risk of developing cervical cancer [6].

Nepal has a cervical cancer incidence rate of 16.4 per 100,000 women, which is nearly four times higher than the WHO’s recommended target of 4 per 100,000 women to eliminate the public health issue of cervical cancer [7]. In FY 2079/80, it was reported that women who were screening for cervical cancer aged 30-49 years had 3.4% and 50+ years had 2.4% had positivity rate [8].

Female community health volunteers (FCHVs) have played a crucial role healthcare service in remote and underserved areas, Nepal. They are selected from their own communities and can bridge the gap between healthcare facilities and the people with those in need. They also lead health of mother groups in communities and facilitate them to bring together like women focusing on safe motherhood; maternal and child health; nutrition; family planning; water, sanitation, and hygiene; community-based health issues and service delivery (Manandhar et al., 2022). They are frontline pillars of community-based health programs who visit every household and advocate healthy behavior among mothers and community people (Khatri, Mishra and Khanal, 2017; Paneru et al., 2023a, 2023b)

The Government of Nepal has acknowledged the significant burden of cervical cancer in the country. In response, the Ministry of Health and Population released the National Guideline for Cervical Cancer Screening and Prevention in 2010. The goal was to screen at least 50% of women in the age group of 30–60 years, which was revised to 70% among 30-49 years in 2017. Only 8.2% of women aged 30–49 years were screened in 2019. However, the screening program did not gain momentum resulting in dismally low coverage owing to implementation difficulties. A recent pilot study was launched to examine the impact of a "community-based intervention on the uptake of cervical cancer screening in a semi-urban area of Pokhara Metropolitan, Nepal [9].Hence, the aim of this research was to identify cervical cancer screening seeking behavior and its associated factors among female community health volunteers in Surkhet district.

## Materials and Methods

### Study design and setting

A cross-sectional study design was employed to identify the cervical cancer screening seeking behavior among FCHVs, utilizing quantitative data collection methods. Data was collected using a structured questionnaire focusing on cervical cancer. The study was conducted in the Surkhet district, Nepal covering an area of 2,489 square kilometers. This is the most populous district in Karnali province which has a total of 4,17,776 population according to the 2021 census. As a central hub for surrounding districts, it has attracted many people for various purposes such as education, business, employment. The district comprises 9 local levels, including 5 municipalities and 4 rural municipalities. There are 990 FCHVs who have crucial roles in various community-based maternal and child programs. Women often feel more comfortable approaching an FCHVs for health information and services. Therefore, FCHV can sensitize women to promote cervical cancer screening seeking behavior effectively.

### Study population and sampling procedure

The study focused on FCHVs aged 30-49 years who were working in specific municipalities and rural municipalities (Birendranagar M, Lekhbesi M, Bheriganga M, Chaukune RM, Baratal RM) within Surkhet district. Simple random sampling was employed to select the municipalities and rural municipalities, with Probability Proportionate to Size (PPS) sampling determining the required sample size from each selected area. Participants were chosen from selected local levels through purposive sampling. Female community health volunteers were excluded from the study if they had joined less than six months ago, had ever been diagnosed with cervical cancer, or had undergone a total hysterectomy.

### Sample size

The sample size was computed by using proportion of 11.3% women of age 30-49 years had undergone cervical cancer screening in Karnali province (Dhimal M. et.al, 2020), 95% confidence interval, 5% margin of error, with finite population of Female health community volunteers in surkhet district N=990. Cochran’s formula (William G. Cochran, 1977) for calculating sample size when the population is infinite:

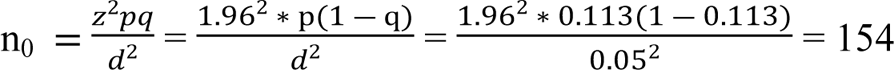

Cochran’s formula for calculating sample size when the population is finite:

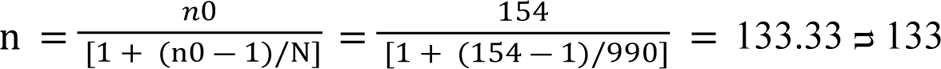

Adding nonresponse rate of 10%,

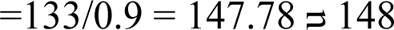

Hence, the required sample will be 148

To calculate the sample size for each municipality and rural municipality, we must calculate k. Here, Population (Ni) = 579 (Total FCHV from the selected municipality and rural municipality) Sample (n) = 148

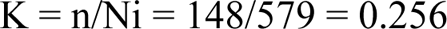

**Table 1:**
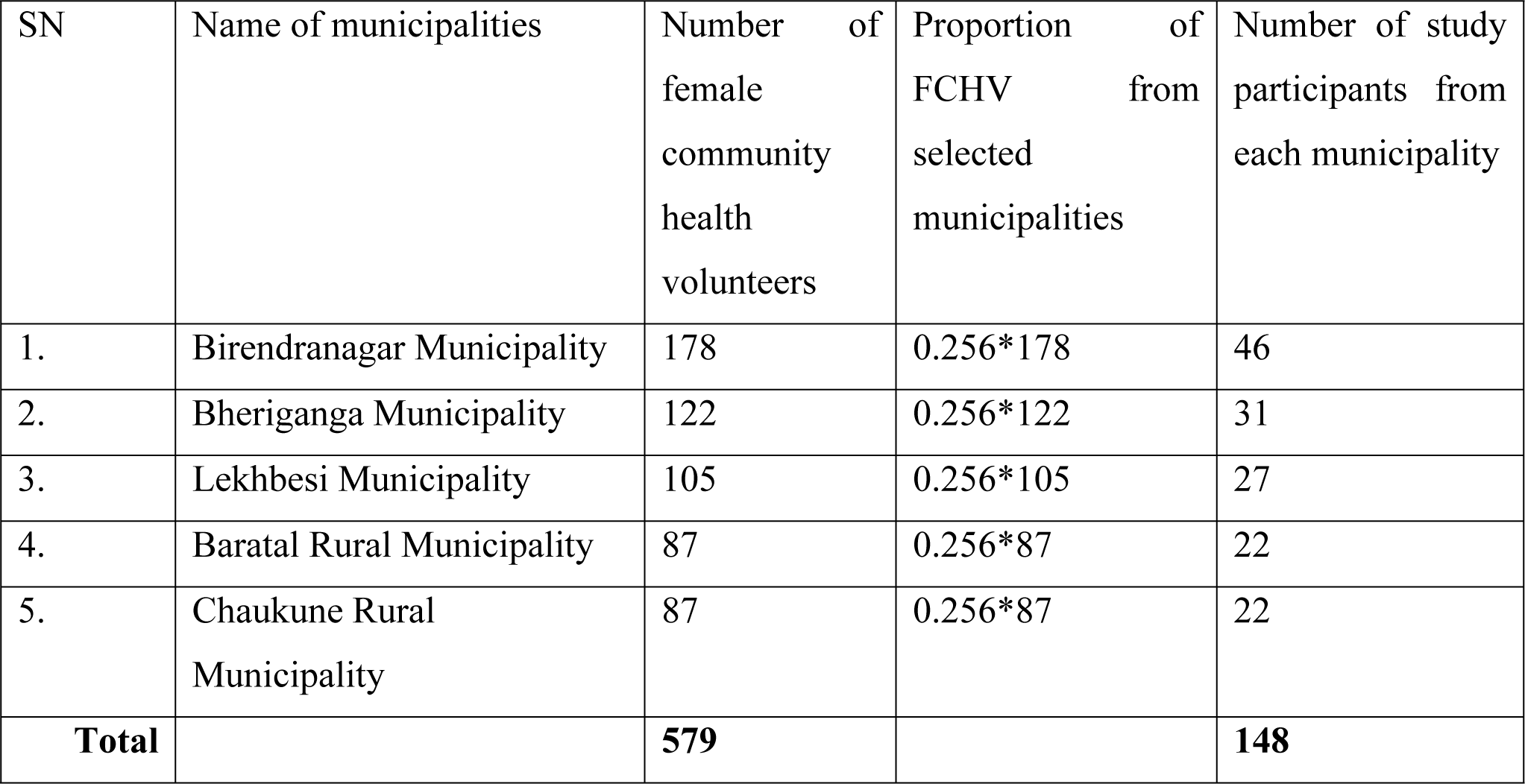
Proportionate allocation random sampling form selected rural/municipalities.

### Study variables

There are eight socio-demographic variables which include age, religion, ethnicity, educational status, marital status, age of marriage, history of miscarriage, age of marriage, monthly income. The occupational factors include duration of profession and the husband’s occupation. The reproductive and obstetric health variables include parity, history of miscarriage/abortion, contraceptive use. Similarly, knowledge about cervical cancer includes, causes/ risk factors, risk age group, preventive measures, screening methods, appropriate age for screening, time difference of screening, symptoms, preventable. The social support variables include peer advice and health personnel advice. Personal behavior includes smoking, the habit of drinking alcohol and multiple sexual partners. The family history of disease consists of HIV/STI and cervical cancer variables. The attitude/perceptions about cervical cancer screening variables include perceived susceptibility, perceived severity, perceived self-efficacy, perceived benefits, perceived barriers.

### Operational definition

The dependent variable, cervical cancer screening seeking behavior refers to FCHV’s utilization of cervical cancer screening facilities. The practice will be assessed by asking the respondent’s action toward screening for premalignant cervical lesions in the past 5 years. Ever screened within the past 5 years were regarded as having good cervical cancer screening seeking behavior. Never screened and screened before 5 years was regarded as having poor cervical cancer screening seeking behavior. It is coded as 1 for good cervical cancer screening seeking behavior and 0 poor cervical cancer screening seeking behavior

Knowledge about cervical cancer screening was evaluated using 27 question items with a “yes” or “no” response. Each correct response was awarded one mark and a wrong response given zero marks. There were 10 questions on cervical cancer causes; 6 on cervical cancer signs and symptoms; 6 questions on cervical cancer prevention and control: 5 questions on others cervical cancer related. The scores from all the 27 items were summed and the mean sum of total scores was calculated. Any participant who scored equal or lesser the mean (≤11 marks) was categorized as having inadequate level of cervical cancer knowledge and participants who scored above the mean (>11 marks) were categorized as having adequate knowledge for cervical cancer. Service availability refers to any method of cervical cancer screening that is available at nearby health facilities.

Service provider refers to the health service providers working at health facilities. Cervical cancer screening methods are pap smear, Visual Inspection with Acetic Acid (VIA), Visual Inspection with Lugol’s Iodine (VILI) and HPV DNA test. VIA is considered as cost-effective method of screening according to WHO for LMICs.

Parity refers to the number of times a woman has given birth to a fetus with a gestational age of 24 weeks or more, regardless of whether the child was born alive or stillborn. It is commonly expressed as a numerical value.

Attitude refers to perception of FCHV towards cervical cancer screening A Likert scale, with a maximum score of five (strongly agree) and a minimum score of one (strongly disagree), was used. All the responses were added up to create a total score, and the mean score was determined as a cut point. Respondents with higher mean scores was deemed to have positive attitudes toward cervical cancer screening, while those with lower scores was deemed to have negative attitude.

### Tools and techniques of data collection

The data was collected through a face-to-face interview administering structured questionnaire. The questionnaires in this study were based on a thorough literature review. The tool was pretested among 10% of total sample among the FCHVs in a similar setting. The internal consistencies of Cronbach’s alpha values for perceived benefit, perceived barrier and perceived severity were 0.7, 0.902 and 0.715 respectively. The questionnaire was prepared in English initially and translated into the Nepali language. The completeness of the data were checked after the completion of the data collection.

### Data processing and analysis

The data was collected using a printed questionnaire and entered in EpiData. Subsequently, the data file was exported to IBM SPSS (Statistical Package for Social Science) version 22 for further analysis. To prevent data loss, the data was stored in a specific folder on a laptop, with backups saved on external hard disk. In quantitative study, the data was transferred from the final excel file to IBM SPSS Statistics 22 for further processing, including cleaning, coding, and analysis. A two-part analysis consisting of descriptive and inferential stages was conducted. Descriptive statistics such as frequency, percentage, mean, and standard deviation were used to present the findings in the form of tables, figures, and text. The Chi-square test, Bi-variate and multivariate logistic regression analyses were performed to identify relationships between variables.

### Ethical considerations

Ethical approval was obtained from the Institutional Review Committee (IRC) of Pokhara University. A written approval to conduct the study was also acquired from the public health service office Surkhet. Written informed consent was taken from each of the participants before the interview and confidentiality and privacy of the participants was maintained. All the participants were informed that there would not be direct benefit or risk to them while participating in this study.

## Results

This chapter includes the descriptive and inferential analysis of various socio-demographic, knowledge, attitude/perceptions, reproductive and obstetric health factors, occupational factors and cervical cancer screening seeking behavior variables. The research instrument was applied to 148 participants, with a 10% non-response rate. The findings of the study are presented below with suitable illustrations.

The socio-demographic characteristics of the respondents are summarized in Table 2. A total of 148 FCHVs, aged 30-49 years with a mean age of 39.7 ±6.6 years, participated in the study. The majority130(87.8%) were Hindu and a third 62(41.9%) were Brahmin/Chettri. 35(23.6%) respondents had no formal education. Most of the respondents 136(91.9%) were married and 120(81.1%) of them had married before the age of 20. More than half of the respondents 90(60.8%) had children ≤2, and 117(79.1%) of women had no history of miscarriage2. 40(37.1%) respondents use hormonal contraceptives which is one of the risk factors of Cervical Cancer. Almost half 73(49.3%) of the respondent had income less than or equal to Rs 20,000 and 77(52%) had health insurance.

**Table 2:** Socio-demographic Characteristics of Respondent.

| Characteristics | Frequency(n=148) | Percentage (%) |
| --- | --- | --- |
| **Age of Respondents** |  |  |
| **Mean age (SD):39.7(±6.6)** |  |  |
| ≤ 34 | 43 | 29.1 |
| 35-39 | 27 | 18.2 |
| 40-44 | 32 | 21.6 |
| 45-49 | 46 | 31.1 |
| **Ethnicity** |  |  |
| Brahmin/Chettri | 62 | 41.9 |
| Dalit | 30 | 20.3 |
| Janajati | 53 | 35.8 |
| Others | 3 | 2.0 |
| **Religion** |  |  |
| Hindu | 130 | 87.8 |
| Buddhist | 8 | 5.4 |
| Christian | 10 | 6.8 |
| **Educational status** |  |  |
| Illiterate | 0 | 0.0 |
| No formal education | 35 | 23.6 |
| Basic level education | 45 | 30.4 |
| Secondary level | 64 | 43.2 |
| Graduate and above | 4 | 2.7 |
| **Marital status** |  |  |
| Married | 136 | 91.9 |
| Divorced | 4 | 2.7 |
| Widowed | 8 | 5.4 |
| **Age at Marriage** |  |  |
| <20 years | 120 | 81.1 |
| ≥20 years | 28 | 18.9 |
| **Parity** |  |  |
| ≤2 | 90 | 60.8 |
| >2 | 58 | 39.2 |
| **Miscarriage** |  |  |
| Yes | 31 | 20.9 |
| No | 117 | 79.1 |
| **Use of family planning method** |  |  |
| Yes | 104 | 70.3 |
| No | 44 | 29.7 |
| **If yes, type of family planning mentioned** |  |  |
| Male sterilization | 29 | 19.6 |
| Female sterilization | 22 | 14.9 |
| IUCD | 2 | 1.4 |
| Implant | 6 | 4.1 |
| Depo | 17 | 11.5 |
| Pills | 15 | 10.1 |
| Condom | 13 | 8.8 |
| None | 44 | 29.7 |
| **Husband's Occupation** |  |  |
| Government employee | 37 | 25.0 |
| Non-government employee | 4 | 2.7 |
| Foreign employment | 33 | 22.3 |
| Agriculture | 33 | 22.3 |
| Daily labour | 23 | 15.5 |
| Business | 12 | 8.1 |
| Unemployed | 2 | 1.4 |
| Retired | 4 | 2.7 |
| **Monthly Family Income Median (Interquartile range): 22000(21500)** |  |  |
| ≤20000 | 73 | 49.3 |
| 20001-40000 | 49 | 33.1 |
| >40000 | 26 | 17.6 |
| **Health insurance** |  |  |
| Yes | 77 | 52.0 |
| No | 71 | 48.0 |

In this study (Table 3), it was found that all 148 (100%) respondents were aware of cervical cancer. The majority, 90 (60.8%), had heard about cervical cancer from health personnel. It was found that 74 (50%) had adequate knowledge, scoring equal to or above the mean (≥11) out of a total of 27. Nearly half, 71 (48%), correctly identified having multiple sexual partners as a risk factor for cervical cancer. Other risk factors mentioned by the respondents included lack of genital hygiene, indicated by 120 (81.1%); early marriage, indicated by 128 (86.5%); HPV virus, indicated by 19 (12.8%); multiple pregnancies, indicated by 82 (55.4%); lack of postnatal care, indicated by 38 (25.7%); prolonged contraceptive use, indicated by 16 (10.8%); hereditary factors, indicated by 25 (16.9%); smoking, indicated by 30 (20.3%); multiple abortions, indicated by 8 (5.4%); and other factors, indicated by 5 (3.5%). More than one fourth, 40 (27%), correctly identified the risk group for cervical cancer. Maintaining genital hygiene was indicated as a major preventive measure by 125 (84.5%), and screening regularly was identified as another preventive measure by 119 (80.4%). More than half, 81 (54.7%), knew the correct age group for cervical cancer screening, but only 32 (21.6%) were aware that VIA is a cost-effective screening method. The majority, 103 (69.6%), identified intra-or post-coital bleeding as a symptom of cervical cancer, 94 (63.5%) reported lower abdominal pain, 91 (61.5%) noted foul-smelling vaginal discharges, and nearly half, 70 (47.3%), thought an itching or burning sensation in the vagina were symptoms of cervical cancer.

**Table 3:**
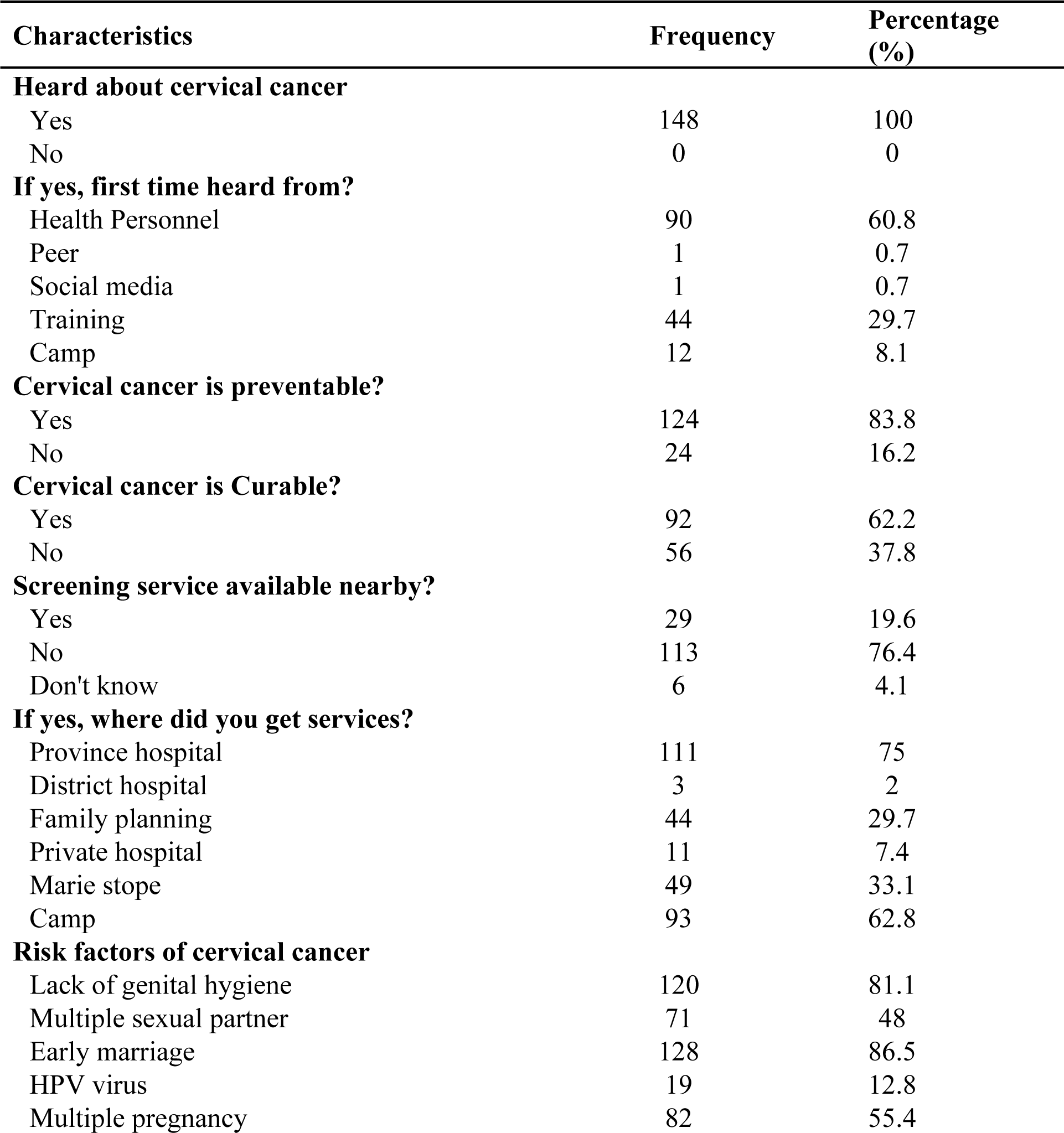
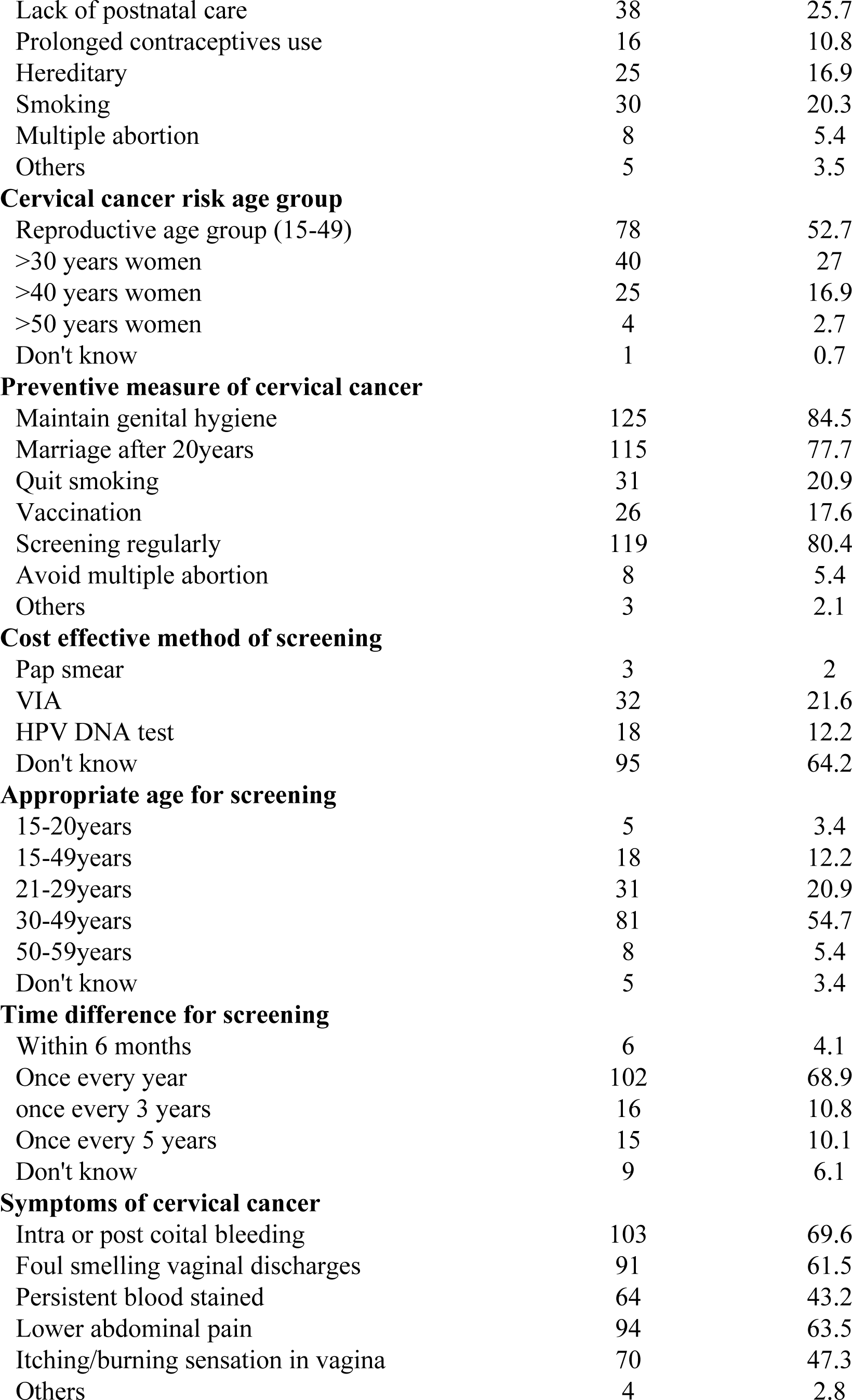
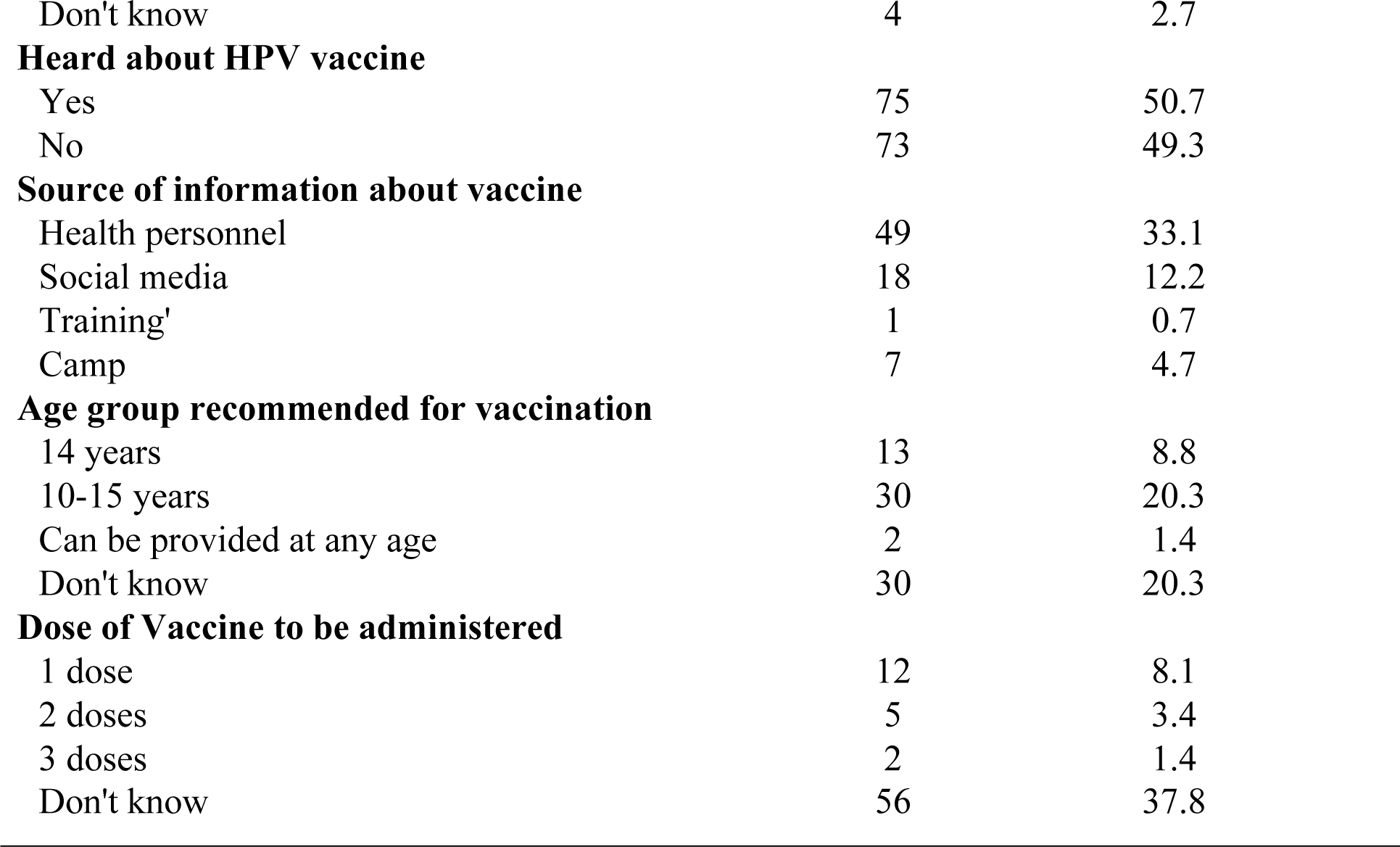
Knowledge regarding cervical cancer (n=148)

Approximately 88 (59.5%) of the participants had undergone a gynecological examination. More than half, 97 (65.5%), had used cervical cancer screening services and the majority 64 (65.97%) had received services from camps. Nearly 39 (40.2%) respondents underwent screening to identify the presence or absence of signs/symptoms, only 9 (9.27%) did screening for the early detection of cervical cancer whereas 30(30.92%) were screened for maintaining good health. Regarding the screening method, only 5 (5.15%) knew that a Pap smear had been performed; 55 (56.7%) were unaware of specific test they underwent and 35(36.08%) knew they had undergone VIA. A total of 66 (44.6%) respondents had been screened once in their lifetime. The primary reasons for not being screened were feeling healthy 35(68.62%) and lack of nearby service 17 (33.33%). Regarding cervical cancer screening, more than half of the participants 90(60.8%) had good screening seeking behavior, while 58(39.2%) had poor screening seeking behavior presented in Table 4.

**Table 4:** Cervical cancer screening seeking behavior.

| Characteristics | Frequency | Percentage (%) |
| --- | --- | --- |
| **Gynecological examination ever** |  |  |
| Yes | 88 | 59.5 |
| No | 60 | 40.5 |
| **Ever screening** |  |  |
| Yes | 97 | 65.5 |
| No | 51 | 34.5 |
| **Service taken from(n=97)** |  |  |
| Government hospital | 9 | 9.27 |
| Private hospital | 11 | 11.34 |
| Family planning | 11 | 11.34 |
| Marie Stope | 2 | 2.06 |
| Camp | 64 | 65.97 |
| **Reason of screening** |  |  |
| Maintenance of good health | 30 | 30.92 |
| Early detection of cervical cancer | 9 | 9.27 |
| Presence/absence of signs and symptom | 39 | 40.2 |
| Since the service is free | 9 | 9.27 |
| To motivate others | 4 | 4.12 |
| Lower abdominal pain | 10 | 10.3 |
| Some other symptoms | 6 | 6.18 |
| **Method used for screening** |  |  |
| Pap smear | 5 | 5.15 |
| VIA | 35 | 36.08 |
| HPV DNA test | 2 | 2.06 |
| Don't know | 55 | 56.7 |
| **Times of screening** |  |  |
| Once | 66 | 68.04 |
| Twice | 18 | 18.55 |
| Thrice or more | 13 | 13.4 |
| **Time of Last screening** |  |  |
| Within 5 years | 90 | 92.78 |
| More than 5 years | 7 | 7.21 |
| **Reason for not screening(n=51)** |  |  |
| Embarrassment | 2 | 3.92 |
| I am healthy | 35 | 68.62 |
| Service is not available nearby | 17 | 17.52 |

Table 5 shows proportion of respondent’s attitude/perceptions about cervical cancer screening.

**Table 5:**
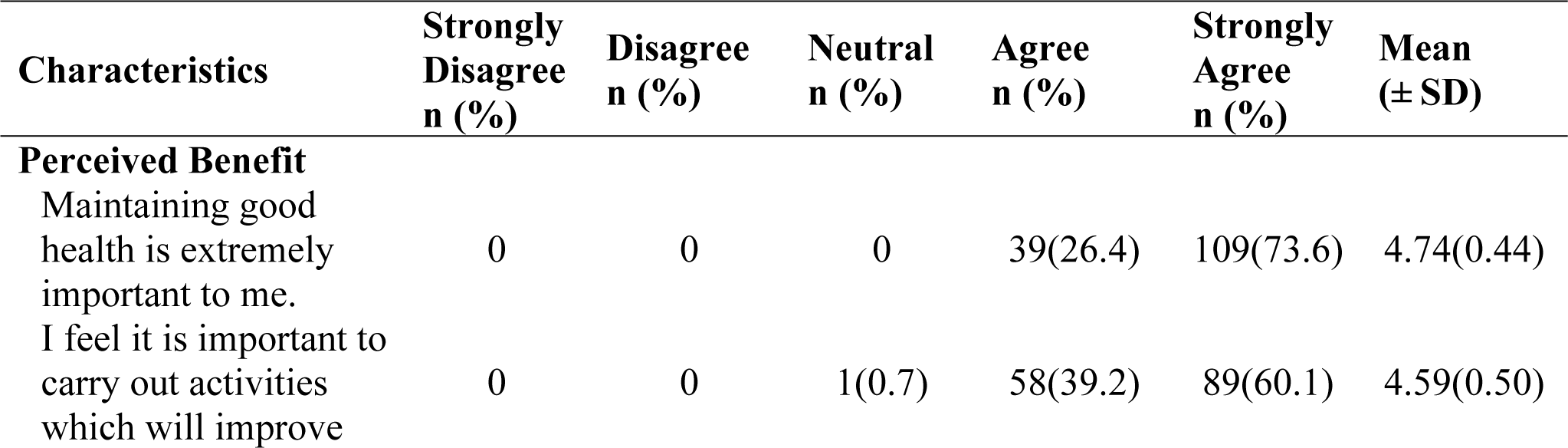
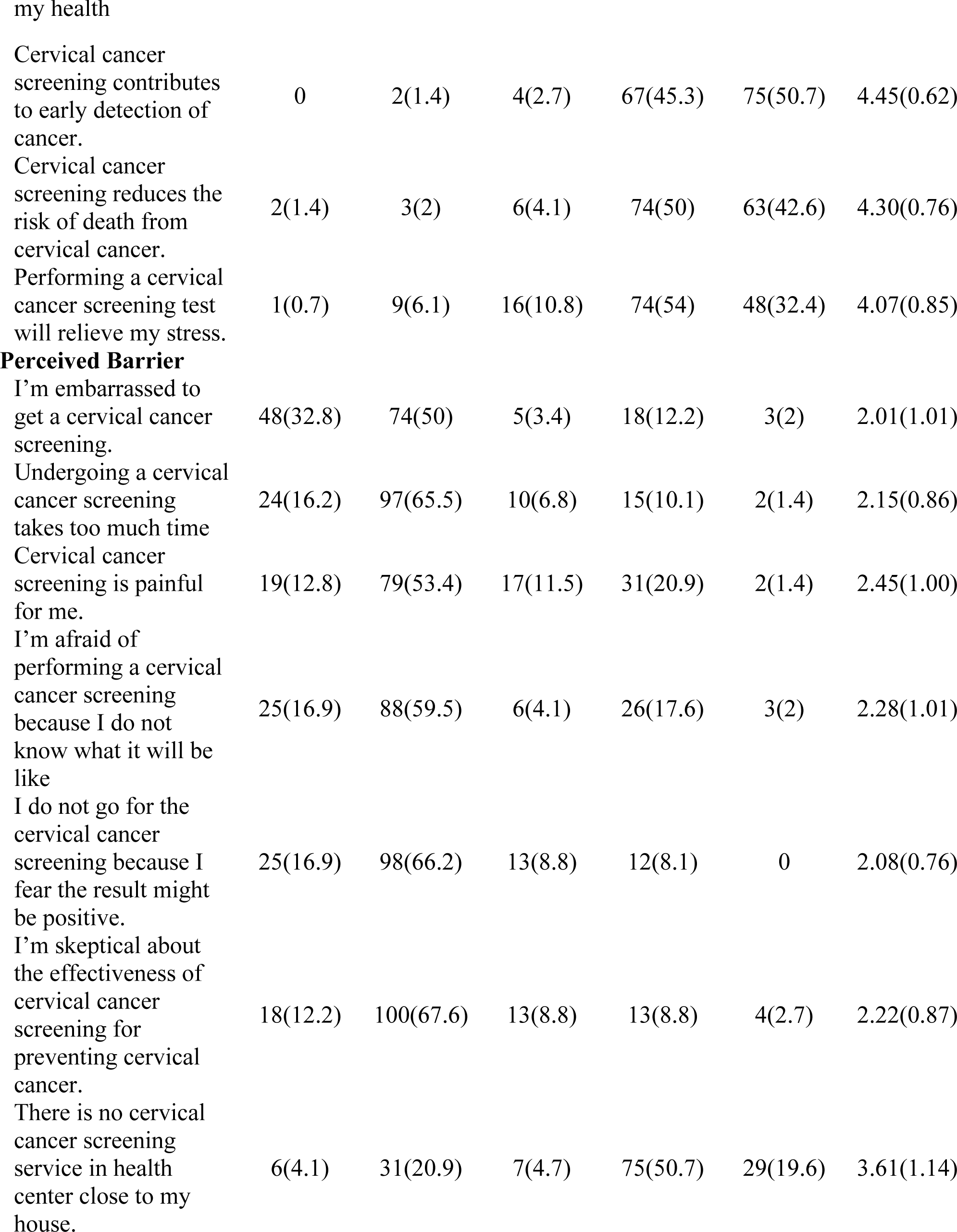
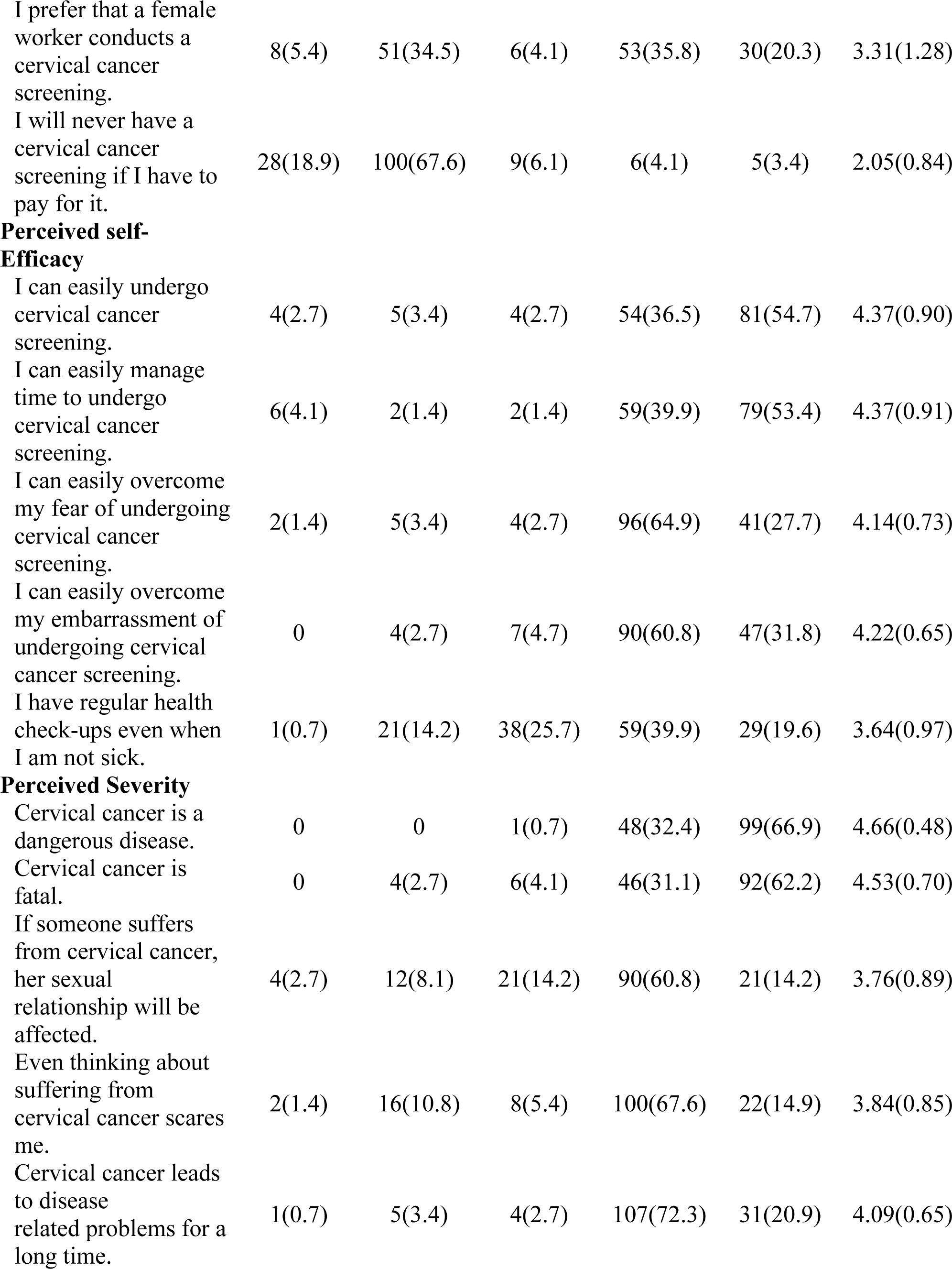
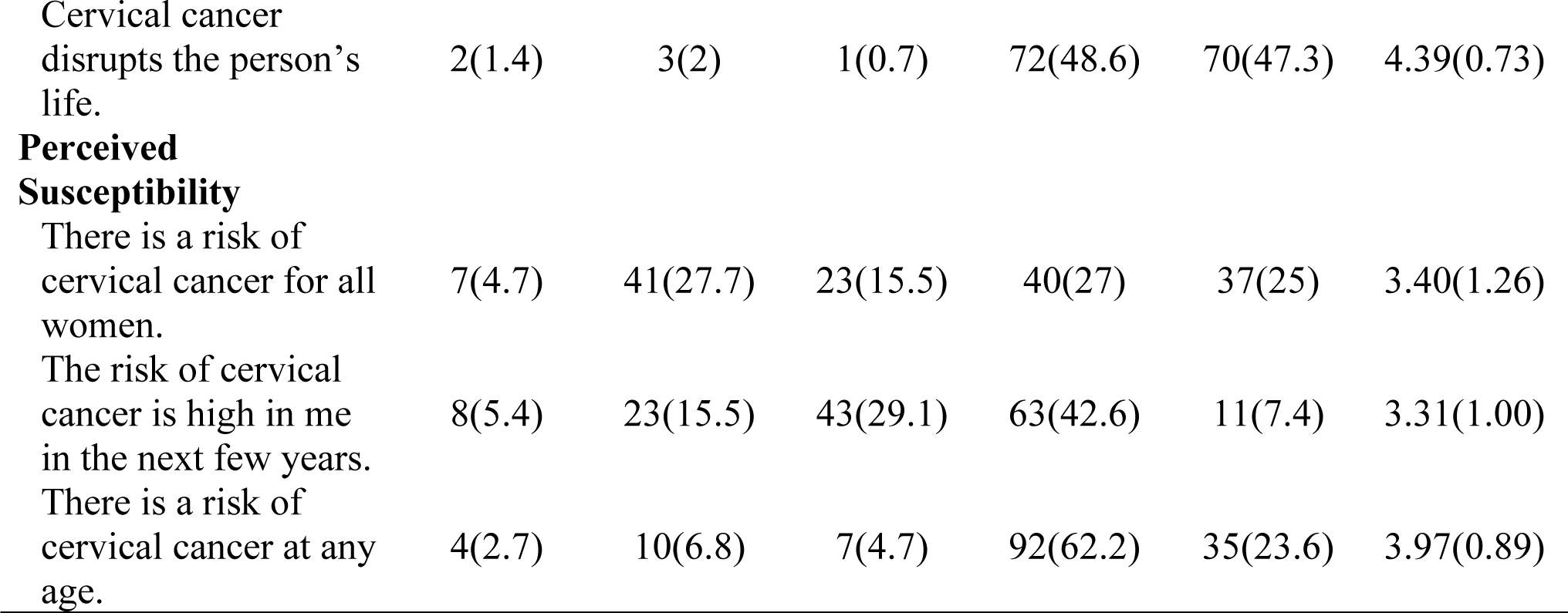
Attitude/perceptions of respondent about cervical cancer screening.

| Characteristics | Strongly Disagree<br>n (%) | Disagree<br>n (%) | Neutral<br>n (%) | Agree<br>n (%) | Strongly Agree<br>n (%) | Mean<br>(± SD) |
| --- | --- | --- | --- | --- | --- | --- |
| **Perceived Benefit** |  |  |  |  |  |  |
| Maintaining good health is extremely important to me. | 0 | 0 | 0 | 39(26.4) | 109(73.6) | 4.74(0.44) |
| I feel it is important to carry out activities which will improve | 0 | 0 | 1(0.7) | 58(39.2) | 89(60.1) | 4.59(0.50) |
| Cervical cancer screening contributes to early detection of cancer. | 0 | 2(1.4) | 4(2.7) | 67(45.3) | 75(50.7) | 4.45(0.62) |
| Cervical cancer screening reduces the risk of death from cervical cancer. | 2(1.4) | 3(2) | 6(4.1) | 74(50) | 63(42.6) | 4.30(0.76) |
| Performing a cervical cancer screening test will relieve my stress. | 1(0.7) | 9(6.1) | 16(10.8) | 74(54) | 48(32.4) | 4.07(0.85) |
| **Perceived Barrier** |  |  |  |  |  |  |
| I'm embarrassed to get a cervical cancer screening. | 48(32.8) | 74(50) | 5(3.4) | 18(12.2) | 3(2) | 2.01(1.01) |
| Undergoing a cervical cancer screening takes too much time | 24(16.2) | 97(65.5) | 10(6.8) | 15(10.1) | 2(1.4) | 2.15(0.86) |
| Cervical cancer screening is painful for me. | 19(12.8) | 79(53.4) | 17(11.5) | 31(20.9) | 2(1.4) | 2.45(1.00) |
| I'm afraid of performing a cervical cancer screening because I do not know what it will be like | 25(16.9) | 88(59.5) | 6(4.1) | 26(17.6) | 3(2) | 2.28(1.01) |
| I do not go for the cervical cancer screening because I fear the result might be positive. | 25(16.9) | 98(66.2) | 13(8.8) | 12(8.1) | 0 | 2.08(0.76) |
| I'm skeptical about the effectiveness of cervical cancer screening for preventing cervical cancer. | 18(12.2) | 100(67.6) | 13(8.8) | 13(8.8) | 4(2.7) | 2.22(0.87) |
| There is no cervical cancer screening service in health center close to my house. | 6(4.1) | 31(20.9) | 7(4.7) | 75(50.7) | 29(19.6) | 3.61(1.14) |
| I prefer that a female worker conducts a cervical cancer screening. | 8(5.4) | 51(34.5) | 6(4.1) | 53(35.8) | 30(20.3) | 3.31(1.28) |
| I will never have a cervical cancer screening if I have to pay for it. | 28(18.9) | 100(67.6) | 9(6.1) | 6(4.1) | 5(3.4) | 2.05(0.84) |
| **Perceived self-Efficacy** |  |  |  |  |  |  |
| I can easily undergo cervical cancer screening. | 4(2.7) | 5(3.4) | 4(2.7) | 54(36.5) | 81(54.7) | 4.37(0.90) |
| I can easily manage time to undergo cervical cancer screening. | 6(4.1) | 2(1.4) | 2(1.4) | 59(39.9) | 79(53.4) | 4.37(0.91) |
| I can easily overcome my fear of undergoing cervical cancer screening. | 2(1.4) | 5(3.4) | 4(2.7) | 96(64.9) | 41(27.7) | 4.14(0.73) |
| I can easily overcome my embarrassment of undergoing cervical cancer screening. | 0 | 4(2.7) | 7(4.7) | 90(60.8) | 47(31.8) | 4.22(0.65) |
| I have regular health check-ups even when I am not sick. | 1(0.7) | 21(14.2) | 38(25.7) | 59(39.9) | 29(19.6) | 3.64(0.97) |
| **Perceived Severity** |  |  |  |  |  |  |
| Cervical cancer is a dangerous disease. | 0 | 0 | 1(0.7) | 48(32.4) | 99(66.9) | 4.66(0.48) |
| Cervical cancer is fatal. | 0 | 4(2.7) | 6(4.1) | 46(31.1) | 92(62.2) | 4.53(0.70) |
| If someone suffers from cervical cancer, her sexual relationship will be affected. | 4(2.7) | 12(8.1) | 21(14.2) | 90(60.8) | 21(14.2) | 3.76(0.89) |
| Even thinking about suffering from cervical cancer scares me. | 2(1.4) | 16(10.8) | 8(5.4) | 100(67.6) | 22(14.9) | 3.84(0.85) |
| Cervical cancer leads to disease related problems for a long time. | 1(0.7) | 5(3.4) | 4(2.7) | 107(72.3) | 31(20.9) | 4.09(0.65) |
| Cervical cancer disrupts the person's life. | 2(1.4) | 3(2) | 1(0.7) | 72(48.6) | 70(47.3) | 4.39(0.73) |
| **Perceived Susceptibility** |  |  |  |  |  |  |
| There is a risk of cervical cancer for all women. | 7(4.7) | 41(27.7) | 23(15.5) | 40(27) | 37(25) | 3.40(1.26) |
| The risk of cervical cancer is high in me in the next few years. | 8(5.4) | 23(15.5) | 43(29.1) | 63(42.6) | 11(7.4) | 3.31(1.00) |
| There is a risk of cervical cancer at any age. | 4(2.7) | 10(6.8) | 7(4.7) | 92(62.2) | 35(23.6) | 3.97(0.89) |

**Table 5:** Attitude/Perceptions of cervical cancer screening.

| Characteristics | Frequency | Percentage % |
| --- | --- | --- |
| **Perceived Benefit** |  |  |
| Positive | 67 | 45.3 |
| Negative | 81 | 54.7 |
| **Perceived Barrier** |  |  |
| Positive | 50 | 33.8 |
| Negative | 98 | 66.2 |
| **Perceived Self efficacy** |  |  |
| Positive | 82 | 55.4 |
| Negative | 66 | 44.6 |
| **Perceived Severity** |  |  |
| Positive | 74 | 50 |
| Negative | 74 | 50 |
| **Perceived Susceptibility** |  |  |
| Positive | 90 | 60.8 |
| Negative | 58 | 39.2 |

Approximately 75(50.7%) respondents had positive attitude of cervical cancer screening. More than half of them 81(54.7%) had negative perceived benefit, 98(66.2%) had negative perceived barrier, 82(55.4%) had positive perceived self-efficacy, 74(50%) had positive perceived severity and 90(60.8%) had positive perceived benefit illustrated by Table 5.

Table 6 demonstrates that half 74(50%) of the respondents possessed adequate knowledge. Additionally, 73(49.3%) respondents exhibited positive attitude and 90(60.8%) of them demonstrated cervical cancer screening seeking behavior.

**Table 6:**
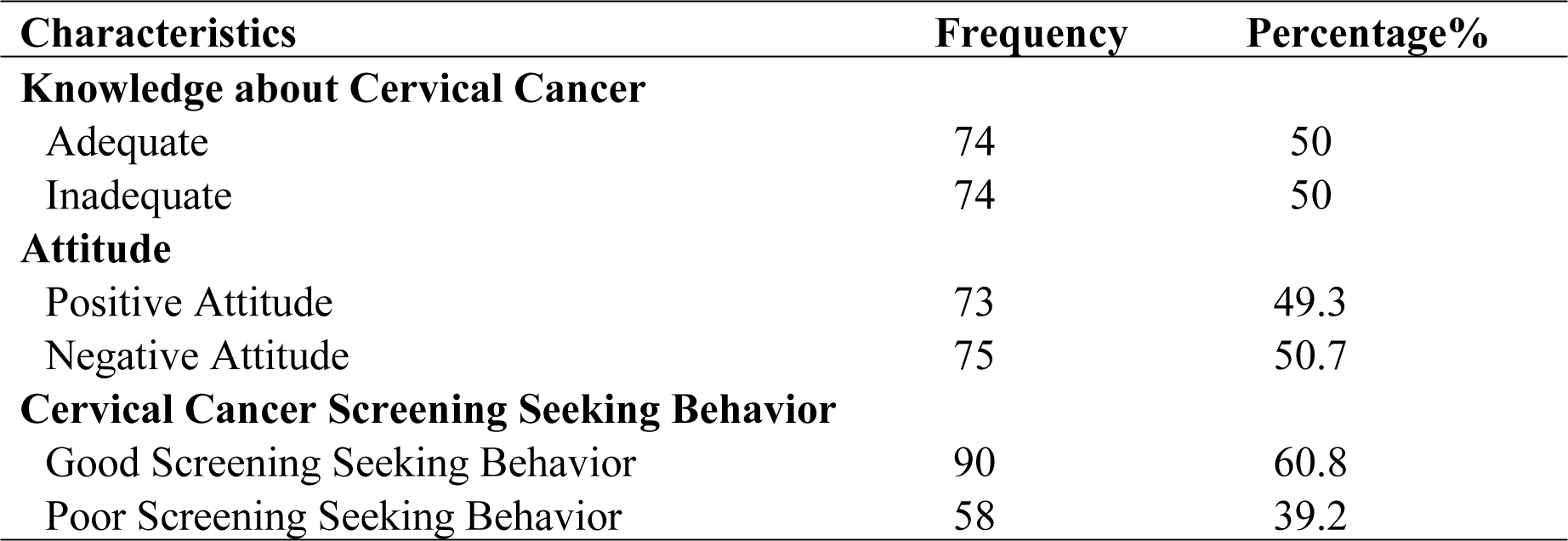
Knowledge, Attitude and Cervical Cancer Screening Seeking Behavior.

| Characteristics | Frequency | Percentage% |
| --- | --- | --- |
| **Knowledge about Cervical Cancer** |  |  |
| Adequate | 74 | 50 |
| Inadequate | 74 | 50 |
| **Attitude** |  |  |
| Positive Attitude | 73 | 49.3 |
| Negative Attitude | 75 | 50.7 |
| **Cervical Cancer Screening Seeking Behavior** |  |  |
| Good Screening Seeking Behavior | 90 | 60.8 |
| Poor Screening Seeking Behavior | 58 | 39.2 |

Table 7 illustrates the cervical cancer screening seeking behavior and its relationship with various socio-demographic characteristics. The major findings indicate that age group (χ2 = 70.909, p-value <0.001) and marital status (χ2 = 15.09, p-value <0.001), were significantly associated with the cervical cancer screening seeking behavior. However, ethnicity, religion, education, age at marriage, number of children, miscarriage, family planning method, family income, health insurance did not have significant association with cervical cancer screening seeking behavior.

**Table 7:**
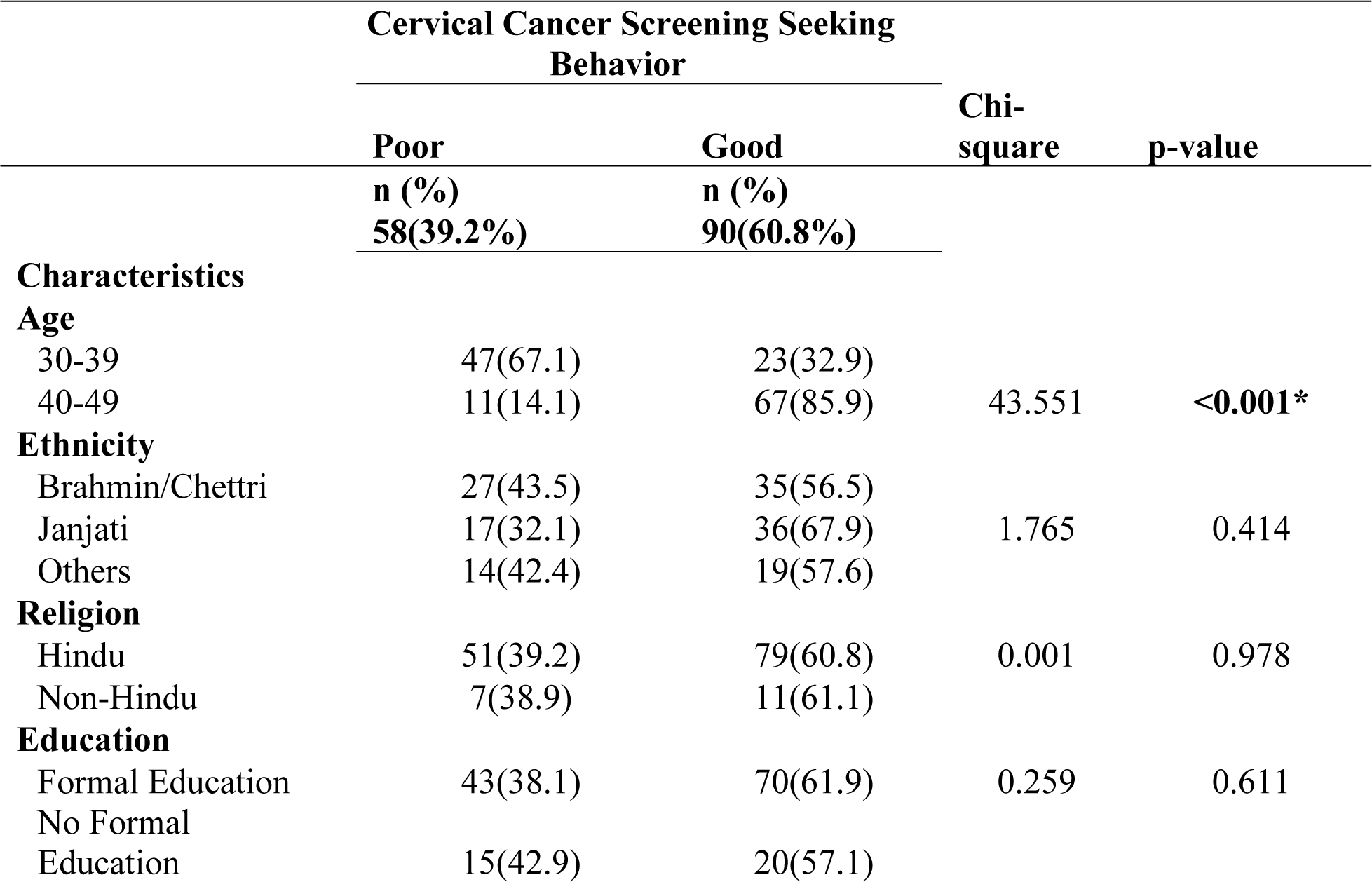
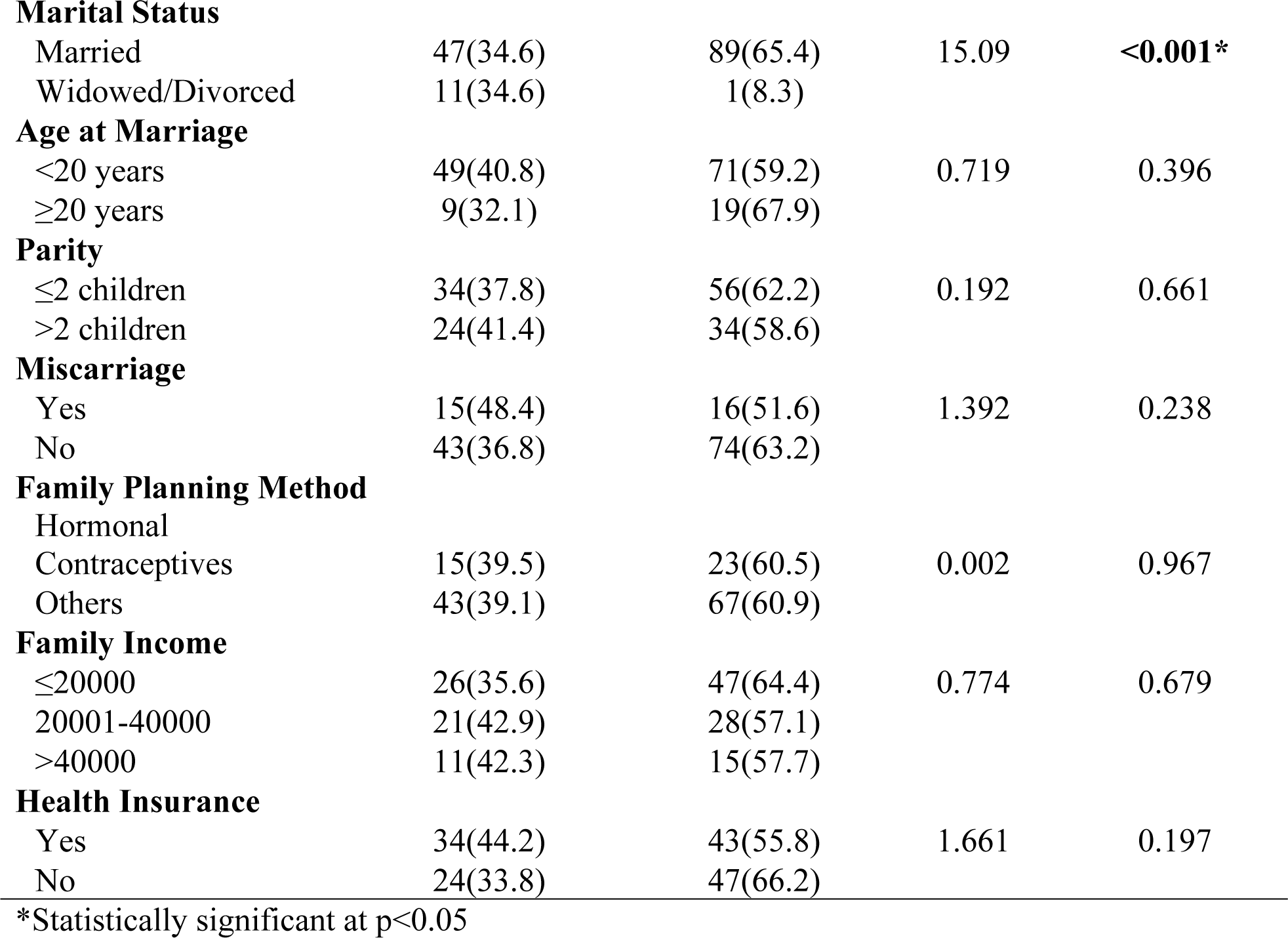
Association of socio-demographic characteristics of respondents with cervical cancer screening seeking behavior.

Table 8 presents the information regarding factors associated with cervical cancer screening seeking behavior. For each variable, the table shows the unadjusted odds ratio (OR), 95 % confidence interval (CI), and p-value, as well as the adjusted OR, 95% CI, and p-value.

**Table 8:** Multivariate analysis of factor associated with cervical cancer screening seeking behavior.

| **Variables** | **Unadjusted OR**<br>**(95% CI)** | **p-Value** | **Adjusted OR**<br>**(95% CI)** | **p-Value** |
| --- | --- | --- | --- | --- |
| **Age Group** |  |  |  |  |
| 30-39 years (Ref) |  |  |  |  |
| 40-49 years | 12.4(5.5-27.9) | <0.001 | 7.2(3.01-17.3) | **<0.001\*** |
| **Marital Status** |  |  |  |  |
| Married | 20.8(2.6-166.2) | 0.004 | 9.2(1.03-83.01) | **0.047\*** |
| Widowed/Divorced (Ref) |  |  |  |  |
| **Work Experience as FCHV** |  |  |  |  |
| Less or equal to 10 years | 2.7(1.3-5.5) | 0.004 | 1.4(0.6-3.4) | 0.405 |
| More than 10 years (Ref) |  |  |  |  |
| **Perceived Benefit** |  |  |  |  |
| Negative | 2.1(1.1-4.2) | 0.024 | 1.3(0.5-3.4) | 0.586 |
| Positive (Ref) |  |  |  |  |
| **Perceived Barrier** |  |  |  |  |
| Negative | 2.8(1.4-5.8) | 0.003 | 1.9(0.7-5.2) | 0.18 |
| Positive (Ref) |  |  |  |  |
| **Attitude** |  |  |  |  |
| Negative | 2.64(1.3-5.2) | 0.005 | 1.6(0.5-4.8) | 0.405 |
| Positive (Ref) |  |  |  |  |

\*Statistically significant at $p < 0.05$ , Ref = Reference

The unadjusted odds ratio revealed that age group 40-49 years were 12.4 times (UOR: 12.4, 95% CI: 5.5-27.9) more likely to screening seeking behavior compared to age group 30-39 years. Likewise, married women 20.8 times more likely to screening seeking behavior compared to widowed/Divorced (UOR: 20.8, 95% CI: 2.6-166.2). Regarding duration of occupation as FCHVs, the odds of screening seeking behavior compared to almost three times higher among FCHVs working less than 10 years (UOR 2.77, 95% CI: 1.3-5.5) compared to those working more than 10 years. Furthermore, participants who had negative perceived benefit were two times more likely to seek screening than those with positive perceived benefit toward cervical cancer screening (UOR 2.1, 95% CI: 1.1-4.2). Regarding, negative perceived barrier was almost two and half times more likely to screen those who had positive perceived barrier towards cervical cancer screening (UOR 2.64, 95% CI: 1.3-5.2).

In adjusted odd ratio, the older age group 40-49 years were 7.2 times (AOR: 7.2, 95% CI: 3.01-17.3) more likely to screen compared to age group 30-39 years. Similarly, Married women were 9.2 times (AOR: 9.2, 95% CI:1.03-83.01) more likely screen compared to widowed/divorced.

## Discussion

### Status of cervical cancer screening seeking behavior

We assessed the status and the associated factors of cervical cancer screening seeking behavior among female community health volunteers. Results show that, despite the existing cost-effective preventive measures, there were only 60.8% of FCHVs have cervical cancer screening seeking behavior. This figure is slightly higher than a similar study in Ethiopia in 2024[10]. However, in a study conducted in Nigeria, 90.6% nurses expressed willingness to be screened but only 24.2% had undergone screening[11].

In contrast a recent study in Ethiopia indicated that only 14.7% female health workforce in public health institutions utilized cervical cancer screening services[12]. Another study conducted in Nepal had 18.3% screening behavior towards cervical cancer[13]. These disparities may result from the Nepalese government’s ongoing initiatives to eradicate cervical cancer. Additionally, the involvement of female community health volunteers (FCHVs) in health campaigns can greatly impact their behavior towards seeking screening.

### Factors affecting cervical cancer screening seeking behavior

The findings of this study revealed that age showed a significant association with cervical cancer screening seeking behavior. Specially, the older age group 40-49 years were 7.2 times (95% CI: 3.01-17.3) more likely to screen compared to age group 30-39 years old. These findings align with a previous four similar study conducted in Ethiopia; Women of advanced age were 13.9 times more likely to utilize cervical cancer screening than younger age (95% CI: 1.40, 136.74) (Hussein et al., 2024). Participants with the age range of 40–49 years were five times more likely to use cervical cancer screening than those whose ages were 30–39 years (95% CI = [2.89, 9.90]) (Belay et al., 2020). Women 40–49 years were likely to utilize cervical cancer screening as compared to women 30–39 years [AOR:3.126(95% CI:1.246, 7.845)][14,15]. The utilization of cervical cancer screening was associated with older age [(AOR) =1.09, CI: 1.07, 8.19][16]. A study of Nepal revealed that women with advanced age were 13 times more likely to perform cervical cancer screening than younger age (95% CI=2.344 73.670)[17]. Older women have a higher risk to developing cervical cancer than younger, however, it is important to consider that if younger women are sexually active. The utilization of screening services is significantly associated with age, mothers over 30 being eight times more likely to use these services compared to those under 30[18]. Women 50 years and older were 4.5 times more likely to have had cervical cancer screening[19].

As women aged, they may become more aware of cervical cancer and the importance of screening due to factors like having children, experiencing routine health checks, or facing age-related health concerns. This could be explained by the fact that as women gets older; their tendency to seek out health care may also increase. The other possible explanation is that when women get older, the risk increases, and they visit medical institutions more frequently. Furthermore, FCHVs are mobilize to community-based intervention with different health training and frequent interactions with health personnel in Nepal, they are more aware of screening.

The findings of this study uncovered that marital status has a significant association with cervical cancer screening-seeking behavior. In this study married women were 9.2 times (95% CI:1.03-83.01) more likely screen compared to widowed/divorced. A study in Nepal resemble with our study, married women were three times more likely to screen for cervical cancer than unmarried women[16]. A study showed married women 10.74 times more likely to uptake cervical cancer screening compared to single women (95% CI = 5.02–22.96)[20]. A study showed association of cervical cancer screening utilization with marital status[AOR:3.41(1.299,8.972)][14]. Another study in Thailand also supported this finding that married/co-inhibiting are 2.26 times more likely undergo cervical cancer screening than those who are single/widowed/divorced (95% CI: 1.58–3.24)[21]. The probable explanation might be the married women perceive themselves as more vulnerable to health issues, motivating them to seek screening.

## Strength and limitation

This study marks the initial research conducted within study area, focusing on assessing the cervical cancer screening seeking behavior and its associated factors among female community health volunteers of Surkhet district.

In this study, data was collected from FCHVs about cervical cancer screening and could not include the general population. The study was limited to the women of age 30-49 years, excludes younger and older women, potentially overlooking important age-specific factors influencing screening behavior. Additionally, recall bias may also affect the study’s findings.

## Conclusion

This study provides a comprehensive analysis of cervical cancer screening behavior among female community health volunteers (FCHVs) in Surkhet, Nepal. Even though there is a proven importance of cervical cancer screening, more than half of the participants exhibit cervical cancer screening seeking behavior. The data reveal significant associations between cervical cancer screening behavior and various socio-demographic factors, particularly age and marital status. Women of advance age are more likely to seek screening compared to younger women. Married women also exhibit higher screening rates than their widowed or divorced counterparts. This suggests that older and married women may have greater health awareness and access to healthcare services.

Additionally, the study highlights the need for targeted interventions to promote cervical cancer screening, particularly among younger and unmarried women, to enhance early detection and improve health outcomes. This underscores the need for targeted interventions that address the specific needs and challenges faced by FCHVs, particularly those newer to the role.

## Data Availability

Not copied

## Acknowledgements

We would like to express our gratitude to all the study participants. In addition, we are grateful towards publich health service office Surkhet, Nepal for providing with administrative support to successfully conduct this study. We would like to acknowledge Faculty members of MPH programme for their encouragement and guidance during the study. We would like to give special thanks to the Institutional Review Committee of Pokhara University for providing ethical approval.

## Author Contribution

Conceptualization: Yamuna Thapa (YT), Bimala Bhatta (BB)

Data Curation: Yamuna Thapa (YT)

Formal Analysis: Yamuna Thapa (YT), Bimala Bhatta (BB)

Investigation: Yamuna Thapa (YT)

Methodology: Yamuna Thapa (YT), Bimala Bhatta (BB)

Writing – Original draft: Yamuna Thapa (YT)

Writing – review and editing: Yamuna Thapa (YT), Bimala Bhatta (BB)

